# Personal protective beliefs and behaviors during the COVID-19 pandemic at a large, multi-campus, public university in Pennsylvania: a cross-sectional survey

**DOI:** 10.1101/2024.09.23.24314227

**Authors:** Jill Stachowski Bell, William Chen, Roxanne E. Kim, Ryan Murphy, Lauren Pomerantz, Prabhani Kuruppumullage Don, Bradley A. Long

## Abstract

**Introduction:** The COVID-19 pandemic impacted many higher education institutions. Understanding the personal protective beliefs and behaviors surrounding public health measures such as facemasking, social distancing, and handwashing helps to inform public health outreach measures.

**Objective:** This study evaluates the beliefs and occurrences of facemasking, social distancing, and handwashing behaviors among people at a multi-campus, public university in Pennsylvania.

**Methods:** From October 5 - November 30, 2021, a 10-minute REDCap survey was available to students, faculty, and staff 18 years of age and older at the Pennsylvania State University (PSU). Recruitment included targeted email, social media, digital advertisements, and university newspapers. 4,231 responses were received. Simple percentage associations between the selected factors and facemasking behaviors as well as beliefs on the origin of the COVID-19 virus were made with IBM SPSS Statistics, version 29.0.2.0 (20). Multivariable logistic regression analyses were performed using R statistical software to identify factors associated with facemasking, social distancing, and handwashing behaviors.

**Results:** Of 3,662 completed surveys, 65.2% of respondents were positive that facemasks prevented the transmission of airborne disease, 49.9% of the respondents stated that they wore facemasks always (27.9%), almost always (26.9%), or often (11.1%). 65.3% believed social distancing prevented transmission of COVID-19, 57.5% reported social distancing while on-campus or out in public (always: 18.8%, almost always: 20.3%, often: 18.4%). The response rate for some level of conscientiously washing hands for the 20 second duration was 87.5% (always: 26.0%, almost always: 31.9%, sometimes: 19.9%, only after being out in public: 4.9%, and only when convenient: 4.8%). Belief in effectiveness of personal protective measures was the strongest predictor of masking and social distancing behaviors. Faculty were more likely than students to report consistent masking, while other behaviors lacked this role-based significance. Social distancing was strongly associated with age and political views, and handwashing was strongly associated with gender.

**Conclusions:** Beliefs in public health and safety measures during the COVID-19 pandemic did not always match the behaviors of individuals surveyed. Understanding barriers to performing behaviors or public health safety may help inform future areas for targeted intervention. Adherence to personal protective behaviors varied across age, gender, institutional role, and political leaning, further emphasizing the importance of targeted public health education.

## Introduction

Facemasks have been worn in healthcare settings for decades, especially during influenza season.^1^ Furthermore, some members of the general public wore facemasks during the 2004-2005 avian flu pandemic.^2^ However, there were no reported human cases of avian influenza A (H5N1) infections within the U.S. during this outbreak, with at least 137 confirmed cases worldwide.^3^ Thus, the scope of the avian flu pandemic affected a much smaller population and was more geographically contained than the COVID-19 pandemic.

During the COVID-19 pandemic, the public was encouraged to wear facemasks to reduce community spread and decrease hospitalizations.^4,5^ Nevertheless, in the United States, facemasking behaviors varied, and pandemonium surrounded facemasks in public settings.^6^ The leading theory was that facemasks had become politicized, explaining the division and divisiveness between those who did not wear them compared to those who did wear them.^7-9^ Studies have shown associations between facemask wearing and misinformation.^10^ Social distancing behavior was also encouraged during the pandemic.^11^ Early studies showed that it helped reduce transmission of the virus, especially at the beginning of the pandemic.^12^ Again, however, the beliefs and behaviors surrounding social distancing became politicized and were subject to misinformation.^13,14^

On the other hand, hand washing was less of a target for misinformation, most likely due to the established history of hand washing as preventing the transmission of disease for centuries.^15^ Yet, to be effective, hand washing is supposed to be performed for a total of 20 seconds; this is less likely of a behavior than expected, handwashing studies have shown that people wash for a shorter duration most of the time.^16^

These behaviors and beliefs were popular topics of debate during the pandemic but were not explored in a large, public university setting in relation to age, gender, and political leaning. The present study evaluated demographic factors that may be associated with differences in mask wearing and different beliefs on the origin of the COVID-19 virus in the fall of 2021, a period when vaccines had become widely available. PSU is one of Pennsylvania’s largest employers, but still represents a higher education community, capturing an interesting subset of the United States population.

## Methods/Subjects

This was a cross-sectional study that took place at all 23 PSU physical campuses, the World Campus, the Penn State Extension, and Penn State Health. Students, faculty, and staff 18 years of age and older were invited to take a REDCap survey between October 5 and November 30 of 2021. The Pennsylvania College of Technology was excluded, as it is only an affiliated institution and not fully part of PSU. The estimated PSU population at the time of the survey was 128,831 (88,914 students, 34,604 faculty and staff, and 5,313 Penn State Health employees).^17-19^ The minimum ideal sample size for this survey was 383 (Confidence Level (Cl): 0.05), as computed by the Qualtrics Sample Size Calculator.^20^

Survey questions were repeatedly tested by medical students, faculty, and staff prior to official launch to receive feedback about question clarity. Participants were recruited by emailing randomly selected 32,792 individuals with a PSU email address, as provided by the Office of the Provost. Additionally, all Penn State Health employees were directly emailed by their Vice President for Human Resources. Digital and print marketing campaigns also took place via campus websites, video boards, print bulletins, the student newspaper, and within university libraries.

This study was carried out in accordance with the institutional review board at the Penn State College of Medicine (reference number 00016908), which deemed this project exempt. Digital, written consent to participate was obtained from participants. They were provided with details and instructions about the research project and asked three qualifying questions. The survey only continued if participants answered “Yes” to all three qualifying questions, which were: consenting to the survey’s instructions, being 18 years of age or older, and being affiliated with the Pennsylvania State University during the study period. If a participant answered “No” to any of these criteria, the survey did not continue. The authors did not have access to information that could identify participants before, during, or after data collection. The REDCap survey incorporated both original research questions and validated questions from previous studies. A simple percentage analysis was conducted using IBM SPSS Statistics, version 29.0.2.0 (20).

Bivariate associations between beliefs and corresponding behaviors were evaluated using Pearson chi-square tests. Multivariable logistic regressions were performed using R statistical software to identify factors associated with adherence to personal protective beliefs. These identifying factors included age, gender, institutional role, and political views, and were self-reported by participants using predefined options within the survey. Three models were constructed for facemasking, social distancing, and handwashing behavior. Within each model, adjusted odds ratios (AOR) with 95% confidence intervals were calculated to estimate the strength of these associations, and statistical significance was defined as p<0.05. Advanced statistical analyses were conducted using R version 4.3.1, and statistical significance was defined as a two-sided p-value < 0.05.

From our prior published analysis of vaccine hesitancy, most participants were between the ages of 26 and 35 (21.0%), women (57.2%), heterosexual (78.0%), and White (77.5%). Most respondents were employees (52.6%).^21^ Of the students who responded, they were primarily undergraduates seeking a bachelor’s degree (63.3%). Additionally, most respondents considered their political views to be liberal (32.9%). Regarding prior COVID-19 infection and vaccination status, a majority never tested positive for COVID-19 (80.3%), have been vaccinated or plan to get vaccinated (80.9%), and received the seasonal influenza vaccine (61.4%).

## Results

For the second analysis of data from this survey, we focused on the beliefs and behaviors of personal protection (facemasking, social distancing, and handwashing) of our survey respondents. Just 65.2% were positive that facemasks prevented the transmission of airborne disease, including COVID-19. (**Table 1**). Conversely, only 49.9% of the respondents stated that they wore facemasks always (27.9%), almost always (26.9%), or often (11.1%).

**Table 1:** Q17: Do you feel that facemasks safely prevent the transmission of airborne diseases, such as the COVID-19 virus?

| Answer | Frequency | Percent |
| --- | --- | --- |
| Yes | 2387 | 65.2 |
| No | 769 | 21 |
| Unsure | 425 | 11.6 |
| Prefer not to answer / Did not answer | 81 | 2.2 |
| **Total** | **3662** | **100** |

Furthermore, 19.0% answered that they wore facemasks only when required. (**Table 2**).

**Table 2:** Q18: How often do you wear a facemask when in public?

| Response | Frequency | Percent |
| --- | --- | --- |
| Always | 1020 | 27.9 |
| Almost always | 985 | 26.9 |
| Often | 405 | 11.1 |
| Sometimes | 188 | 5.1 |
| Once in a while | 82 | 2.2 |
| Almost never | 99 | 2.7 |
| Only when required | 695 | 19 |
| I haven't worn a mask since vaccines have become widely available | 45 | 1.2 |
| I haven't worn a mask since the CDC guidelines first removed the need to wear one in public | 64 | 1.7 |
| Prefer not to answer / Did not answer | 79 | 2.1 |
| **Total** | **3662** | **100** |

A multivariable logistic regression analysis was completed to examine factors independently associated with consistent facemasking. Belief that facemasks prevent COVID-19 was the strongest predictor of masking behavior, as evident by an adjusted odds ratio of 5.93, 95% Cl: 4.97-7.09, p<0.001; **Table 3**). In addition to belief, several demographic factors were also associated with masking behavior. Women (AOR = 1.43, 95% Cl: 1.20-1.70) and “Other” (AOR = 2.52, 95% Cl: 1.06-6.21) gender-identifying individuals were more likely to report consistent masking compared to men (**Table 3**). People identifying as far left (AOR = 8.74, 95% Cl: 6.69-11.5), liberal (AOR = 5.04, 95% Cl: 3.44-7.50), or middle-of-the-road (AOR = 2.24, 95% Cl: 1.79-2.79) in political view had higher odds of consistent masking behavior as opposed to those identifying as far right (**Table 3**) . Faculty were significantly more likely than students to report consistent masking (AOR= 1.53, 95% Cl: 1.12-2.09, p=0.007), while staff did not differ from students (AOR = 1.06, 95% Cl: 0.85-1.31, p=0.605; **Table 3**). Overall, consistent masking behavior was strongly associated with belief in effectiveness in COVID-19 prevention, followed by political views and certain demographic factors.

**Table 3:** Multivariable logistic regression of factors associated with consistent facemask use.

| Predictor | Adjusted Odds Ratio (AOR) | 95% CI | p-value |
| --- | --- | --- | --- |
| **Belief that facemasks prevent COVID-19 transmission** | **5.93** | **4.97–7.09** | **<0.001** |
| **Age (ref: 18–25)** |  |  |  |
| 26–35 | 0.57 | 0.43–0.76 | <0.001 |
| 36–45 | 0.67 | 0.51–0.89 | 0.006 |
| 46–55 | 0.74 | 0.55–0.99 | 0.045 |
| 56–65 | 0.83 | 0.61–1.12 | 0.213 |
| >65 | 1.12 | 0.81–1.55 | 0.5 |
| Prefer not to answer | 2.86 | 1.36–6.47 | 0.007 |
| **Gender (ref: Man)** |  |  |  |
| Woman | 1.43 | 1.20–1.70 | <0.001 |
| Other | 2.52 | 1.06–6.21 | 0.039 |
| Prefer not to answer | 0.58 | 0.30–1.10 | 0.101 |
| **Role (ref: Student)** |  |  |  |
| Faculty | 1.53 | 1.12–2.09 | 0.007 |
| Staff | 1.06 | 0.85–1.31 | 0.605 |
| **Political views (ref: Far right)** |  |  |  |
| Far left | 8.74 | 6.69–11.5 | <0.001 |
| Liberal | 5.04 | 3.44–7.50 | <0.001 |
| Middle-of-the-road | 2.24 | 1.79–2.79 | <0.001 |
| Right | 0.66 | 0.32–1.28 | 0.236 |

Next, beliefs and behaviors related to social distancing were analyzed. Just 65.3% believed that social distancing prevented transmission of COVID-19 (**Table 4**). 57.5% of the respondents reported that they commonly social distanced while on campus or out in public (always:18.8%, almost always: 20.3%, often: 18.4% (**Table 5**).

**Table 4:**
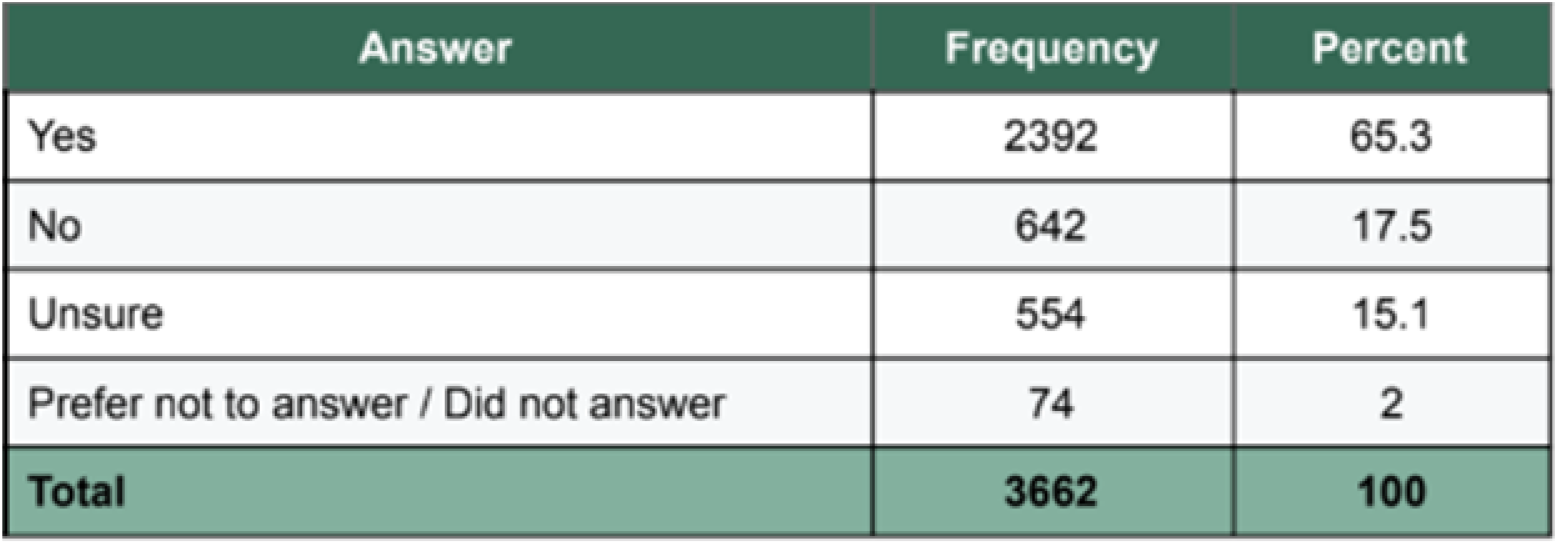
Q19: Do you feel that social distancing safely prevents the transmission of the COVID-19 virus?

**Table 5:**
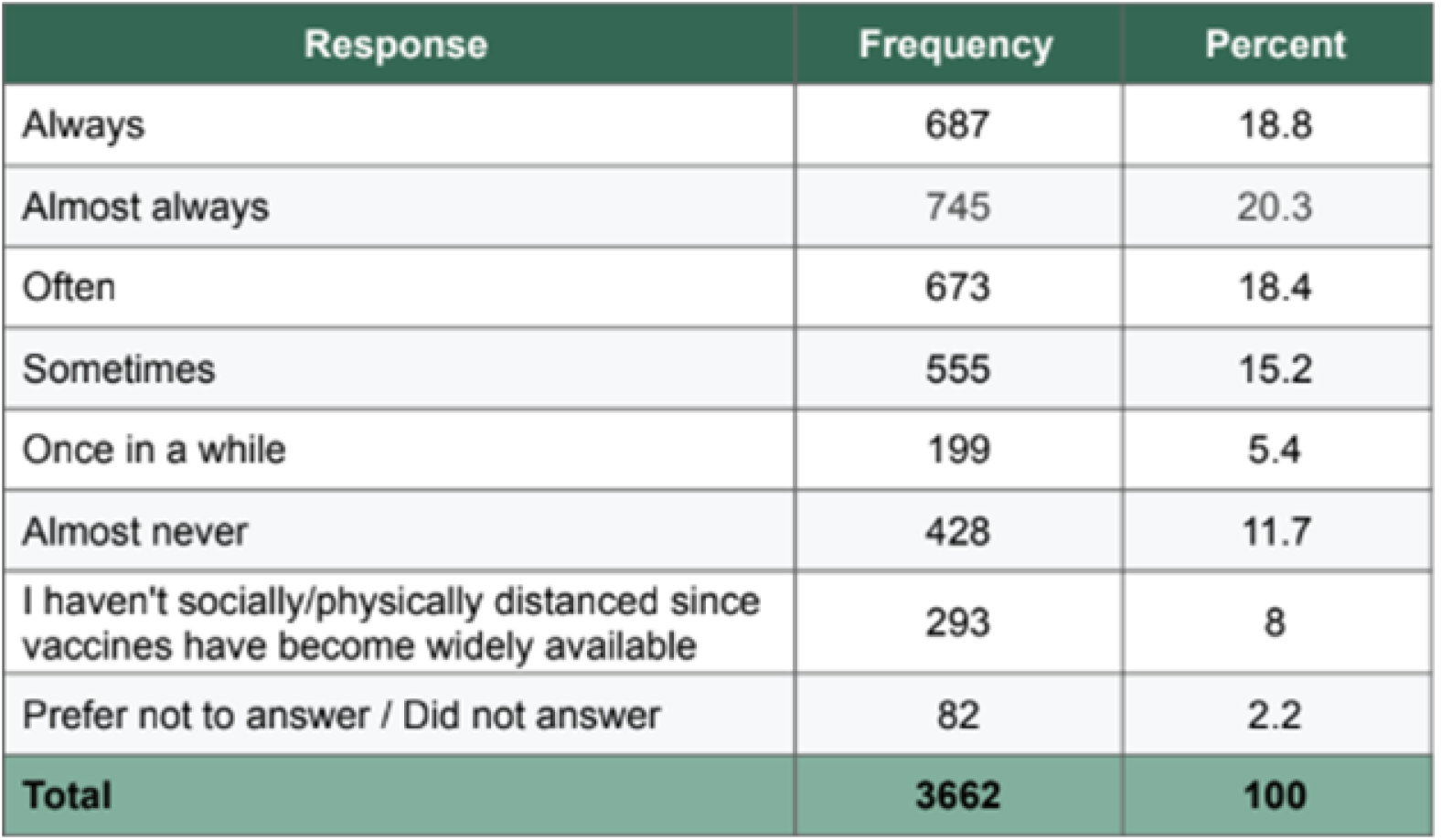
Q20: Do you attempt to social/physical distance when on campus or out in public?

According to multivariable logistic regression, belief that social distancing prevents COVID-19 transmission was strongly associated with social distancing behavior (AOR=4.42, 95% Cl: 3.75-5.22, p<0.001; **Table 6**). Age group demonstrated a significant incline, with increasing age associated with higher odds of social distancing. For example, the adjusted odds ratio for age group 26-35 was 2.42 with p-value<0.001 and it rose to AOR = 9.68 with p<0.001 in the >65 age group (**Table 6**). Women were significantly more likely than men to report consistent distancing (AOR = 1.35, 95% Cl = 1.15-1.58, p<0.001; **Table 6**). Similarly to facemasking behavior, individuals identifying as far left, liberal, or middle-of-the-road reported distancing significantly more than those identifying as far right (**Table 6**).

**Table 6:** Multivariable logistic regression of factors associated with consistent social distancing behavior.

| Predictor | Adjusted Odds Ratio (AOR) | 95% CI | p-value |
| --- | --- | --- | --- |
| **Belief that social distancing prevents COVID-19 transmission** | **4.42** | **3.75–5.22** | **<0.001** |
| **Age (ref: 18–25)** |  |  |  |
| 26–35 | 2.42 | 1.87–3.12 | <0.001 |
| 36–45 | 2.79 | 2.13–3.67 | <0.001 |
| 46–55 | 2.73 | 2.08–3.60 | <0.001 |
| 56–65 | 3.95 | 2.94–5.34 | <0.001 |
| >65 | 9.68 | 5.01–19.9 | <0.001 |
| Prefer not to answer | 7.36 | 2.44–23.0 | <0.001 |
| **Gender (ref: Man)** |  |  |  |
| Woman | 1.35 | 1.15–1.58 | <0.001 |
| Other | 1.07 | 0.53–2.19 | 0.86 |
| Prefer not to answer | 0.63 | 0.33–1.18 | 0.16 |
| **Role (ref: Student)** |  |  |  |
| Faculty | 1.02 | 0.77–1.36 | 0.881 |
| Staff | 0.91 | 0.74–1.12 | 0.369 |
| **Political views (ref: Far right)** |  |  |  |
| Far left | 6.16 | 4.35–8.83 | <0.001 |
| Liberal | 5.06 | 4.03–6.38 | <0.001 |
| Middle-of-the-road | 2.38 | 1.91–2.97 | <0.001 |
| Right | 0.94 | 0.45–1.84 | 0.86 |

The respondents were also asked if they washed their hands for at least 20 seconds. The response rate for some level of conscientiously washing hands for the 20-second duration was 87.5% (always: 26.0%, almost always: 31.9%, sometimes: 19.9%, only after being out in public: 4.9%, and only when convenient: 4.8%. 2.3% only used hand sanitizer. (**Table 7**).

**Table 7:** Q21: It is still recommended that after coming into contact with a potentially unclean surface that you wash your hands with soap and water for at least 20 seconds. How often do you wash your hands for at least 20 seconds?

| Response | Frequency | Percent |
| --- | --- | --- |
| Always | 952 | 26 |
| Almost always | 1170 | 31.9 |
| Sometimes | 728 | 19.9 |
| Only after being out in public | 179 | 4.9 |
| Only when convenient | 175 | 4.8 |
| Rarely | 180 | 4.9 |
| Almost never | 79 | 2.2 |
| I have not followed this practice since vaccines have become widely available | 39 | 1.1 |
| I only use hand sanitizer | 85 | 2.3 |
| Prefer not to answer / Did not answer | 75 | 2 |
| **Total** | **3662** | **100** |

Being that handwashing safety is a common part of early childhood education, in addition to keeping the survey length to a reasonable time to complete, no question was added regarding handwashing safety beliefs. A multivariable logistic regression found that out of the demographic factors examined, age and gender had the strongest associations to handwashing behavior (**Table 8**). Individuals in age groups 56-65 (AOR = 1.42, 95% Cl: 1.12-1.79, p = 0.004) and older than 65 (AOR = 1.34, 95% Cl: 1.04-1.72, p = .022) demonstrated significantly higher odds of reported consistent handwashing than individuals aged 18-25 (**Table 8**). Women were significantly more likely than men to report consistent handwashing (AOR = 1.53, 95% Cl: 1.34-1.76, p<0.001; **Table 8**).

**Table 8:** Multivariable logistic regression of factors associated with consistent handwashing behavior.

| Predictor | Adjusted Odds Ratio (AOR) | 95% CI | p-value |
| --- | --- | --- | --- |
| **Age (ref: 18–25)** |  |  |  |
| 26–35 | 0.92 | 0.74–1.14 | 0.447 |
| 36–45 | 1.11 | 0.89–1.37 | 0.354 |
| 46–55 | 1.13 | 0.90–1.42 | 0.288 |
| 56–65 | 1.42 | 1.12–1.79 | 0.004 |
| >65 | 1.34 | 1.04–1.72 | 0.022 |
| Prefer not to answer | 1.59 | 0.98–2.63 | 0.064 |
| **Gender (ref: Man)** |  |  |  |
| Woman | 1.53 | 1.34–1.76 | <0.001 |
| Other | 1.08 | 0.63–1.86 | 0.777 |
| Prefer not to answer | 0.52 | 0.28–0.94 | 0.034 |
| **Role (ref: Student)** |  |  |  |
| Faculty | 0.76 | 0.58–1.01 | 0.055 |
| Staff | 0.96 | 0.78–1.17 | 0.662 |
| **Political views (ref: Far right)** |  |  |  |
| Far left | 1.17 | 0.88–1.55 | 0.284 |
| Liberal | 0.73 | 0.44–1.21 | 0.229 |
| Middle-of-the-road | 1.32 | 1.10–1.59 | 0.003 |
| Right | 1.15 | 0.96–1.39 | 0.14 |
| Prefer not to answer | 1.13 | 0.86–1.48 | 0.386 |

Political views, with the exception of middle-of-the-road individuals (AOR = 1.32, 95% Cl: 1.10-1.59, p=0.003), showed weak and inconsistent associations with handwashing in contrast to facemasking (**Table 3**) and social distancing behaviors (**Table 6**). Institutional role also appeared to have an insignificant influence on handwashing behavior (**Table 8**)..

## Discussion

The results indicate that in the case of COVID-19 protective measures, even when people believed in the correct information surrounding public health safety behaviors, or at least stated that they believed in them, they did not actually perform the behaviors. This was especially true in the cases of face masking and social distancing. 65% of respondents felt that face masking did prevent transmission of COVID-19, yet only roughly 28% and 27% of respondents always or almost always wore them. Similarly, 65% of respondents felt that social distancing helped to reduce the spread of COVID-19, but only 19% and 20% of respondents almost or almost always socially distanced themselves. It could be implied that individuals do believe in the power of handwashing, and this can be extrapolated to include the handwashing behaviors: if individuals do believe in the power of handwashing, there should be more “always” or “almost always” responses seen but only 26% and 32% do handwash for at least 20 seconds.

The results indicated that there are not only challenges to encouraging people to believe that certain public health measures are protective for their health, but also to encouraging them to act in a manner consistent with those beliefs. During the COVID-19 pandemic, theories are that the politicization of behaviors and the spread of misinformation in the pandemic led to poor outcomes.^22^ Yet, it is possible that beliefs were present, but the motivations of the behaviors were misaligned with a common goal of health and safety of the public.

Multivariable regression analysis was used to identify factors associated with protective behaviors. These analyses demonstrated that adherence to protective behaviors varied across demographics, political views, and institutional roles. Reported belief in the effectiveness of protective measures was one of the strongest predictors of behavior for facemasking and social distancing. Additionally, political views were strongly associated with masking and distancing behaviors, supporting previous work that these behaviors were shaped by sociopolitical influences. ^7-9, 22^ In this study, the strongest predictor associations outside of belief were between facemasking and institutional role and beliefs, social distancing and age and political views, and handwashing and gender. These findings suggest that there are likely differences in how various groups interpret public health information and practice personal protective beliefs or lack thereof.

This study has several limitations, such as being a single institution study and reliance on self-reporting for major endpoints of the study - including the personal protection specific questions. Participants may have had difficulty selecting survey responses if the available answers were unclear to them, and respondents may have made some misinformed but well-meaning answers. Furthermore, the first study utilized a graduate-level statistics student to conduct the analysis using R software.^21^ The statistical analysis for this study was conducted in two parts, separated by a full academic year. The percentage analyses were conducted first using SPSS statistical software with individuals possessing limited statistical experience, a 4th-year medical student and a medical librarian. The multivariable regression analyses were conducted a year later by two 1st-year medical students with significant prior experience using R software.

Despite the limitations, this survey study showed that belief in public health and safety measures during the COVID-19 pandemic did not always match the behaviors of individuals. This has key implications for future public health information campaigns, where convincing individuals in the validity of the science and the behaviors may not be enough to get them to be invested in making a change for their own health and safety or the health and safety of society.^22^ A further step would be to understand the barriers to performing the behaviors and any key factors negatively influencing their behaviors.

## Conclusion

Beliefs in public health and safety measures during the COVID-19 pandemic did not always match the behaviors of individuals surveyed. Understanding barriers to performing behaviors or public health safety may help inform future areas for targeted intervention. Further future analysis may be required to determine why individual beliefs and behaviors did not necessarily match.

## Data Availability

All personally non-identifiable data produced are available online at Mendeley Data.

https://data.mendeley.com/datasets/vvnsj7n56t/2

## Supporting Information

Long B, Murphy R, Pomerantz L, Kuruppumullage Don P. Pennsylvania State University Fall 2021 COVID-19 Infodemic Survey (ver 2). Mendeley Data. Published Sep 27, 2023. Accessed Apr 28, 2026. doi:10.17632/vvnsj7n56t.2

